# Aligning Reinforcement Learning with Clinical Practice for Safe Decision Support in Pediatric Sepsis

**DOI:** 10.64898/2026.07.20.26358476

**Authors:** Fernanda Bueso Medina, Richard Wardle, Petru Manescu, Joshua Spear, Samiran Ray, Mark Peters

## Abstract

Offline reinforcement learning (RL) has emerged as a promising framework for clinical decision support in sepsis, yet most existing studies focus exclusively on adult populations, leaving pediatric care largely unexplored despite important physiological and treatment differences. In this work, we develop offline RL policies for pediatric sepsis management in the Pediatric Intensive Care Unit (PICU) using a retrospective cohort of 2,229 episodes from Great Ormond Street Hospital (GOSH), formalized as finite-horizon Markov Decision Process (MDP) with joint intravenous fluid and vasopressor actions. To better capture pediatric organ dysfunction dynamics, we incorporate Phoenix-8, a recently proposed pediatric sepsis severity score, as an intermediate reward shaping signal in addition to terminal 90-day mortality. We systematically vary the time-step size (4, 8, and 12 hours) and reward structure (terminal 90-day mortality, with and without Phoenix-8–based intermediate shaping), and compare Double Deep Q-Networks (DDQN), Conservative Q-Learning (CQL), and a behavior cloning (BC) model of clinician practice. CQL consistently exhibits stable learning dynamics and favorable Fitted Q Evaluation estimates, while DDQN is prone to overestimation and instability, particularly at finer temporal resolutions and with dense rewards. CQL policies achieve high action-level agreement with historical clinician decisions for both fluids and vasopressors and reproduce clinically plausible escalation patterns across sepsis severity strata, whereas DDQN policies diverge more frequently toward implausible dosing. Temporal aggregation emerges as a key regularizer: moving from 4-hour to 8-hour bins shortens horizons, smooths reward noise, and improves stability without erasing clinically meaningful dynamics, with 8-hour binning providing the best trade-off between policy performance and granularity. Our findings highlight time-step size as a core design choice in offline RL for healthcare and provide empirical evidence that alternatives beyond the conventional 4-hour setup can enhance stability and safety while preserving clinical interpretability.

**Author summary:** We studied how artificial intelligence might support treatment decisions for children with sepsis in intensive care. Sepsis is a serious condition that can worsen quickly, and clinicians often need to decide how much fluid or blood pressure support to give over time. Although artificial intelligence has been studied for adult sepsis, much less is known about how these methods perform in children, whose illness patterns and treatment needs can differ in important ways. Using records from 2,229 pediatric intensive care admissions, we tested whether a learning system could identify treatment strategies that were both stable and consistent with real clinical practice. We found that some modeling choices had a major effect on the safety and credibility of the recommendations. In particular, grouping data into 8-hour intervals produced more reliable results than the more commonly used 4-hour approach, while still preserving meaningful changes in illness severity. These results suggest that safe and useful decision-support tools for pediatric sepsis depend not only on the choice of the right algorithm, but also on careful design choices. We hope this work helps guide future research toward more trustworthy clinical artificial intelligence.

## Introduction

Sepsis is a life-threatening condition characterized by organ dysfunction resulting from a dysregulated immune response to infection [1, 2]. Despite advances in critical care, current approaches to sepsis management largely rely on standardized guidelines, the experience of the clinician, and scoring systems. While valuable, these methods are often limited by the complexity of patient trajectories and the inherent variability of real world healthcare data. This has sparked a growing interest in data-driven techniques, which have the ability to identify subtle patterns in historical patient data and guide personal treatment strategies. In this context, artificial intelligence (AI) methods have emerged as promising tools to support clinical decision-making in critical care.

Sepsis provides a compelling case study for reinforcement learning because management requires sequential decisions under uncertainty, and timely interventions based on dynamically observable patient states are believed to play a critical role in altering the course of illness. Clinical care evolves over time through repeated interventions such as fluid resuscitation and vasopressor administration, while patient states change dynamically in response to both disease progression and therapy. Reinforcement learning (RL), which aims to learn optimal decision policies from interactions with an environment, provides a natural framework for modeling this process. In many clinical settings, including sepsis management, there is often no single universally optimal treatment strategy supported by randomized evidence. As a result, clinical decision-making varies across practitioners and institutions, creating an opportunity for RL methods to learn effective treatment policies from observed practice patterns. Because real-time experimentation is infeasible in clinical practice, offline RL approaches that learn exclusively from retrospective datasets have become particularly attractive in healthcare applications [3–6]. These methods are capable of handling the high dimensional nature of ICU data, which includes demographics, vital signs, and laboratory tests recorded at irregular intervals [3, 5]. By learning directly from outcomes rather than relying solely on clinical heuristics, RL has the potential to refine or even challenge existing guidelines such as the Surviving Sepsis Campaign, particularly in domains where evidence remains limited (e.g., optimal fluid resuscitation volumes) [6].

Early work applying RL to sepsis care focused primarily on adult intensive care populations. The landmark study by Komorowski, et al. [4] introduced the “AI Clinician,” an offline RL model trained on the adult ICU MIMIC-III dataset. The AI Clinician learned treatment policies by analyzing thousands of patient trajectories, identifying sequences of interventions that were associated with improved outcomes. For example, it learned how the timing and dosage of fluids and vasopressors influenced long term survival, rather than focusing only on short term physiological targets. By framing sepsis care as a sequential decision making problem, the study demonstrated the potential of RL to move beyond static prediction models toward adaptive, patient-specific treatment strategies. Subsequent studies have explored how RL can support personalized treatment recommendations, adapting interventions to account for the heterogeneity of sepsis presentations across individual patients [3, 5]. Technical advances have also tackled the issue of sparse and delayed rewards in adult cohorts by incorporating intermediate scoring systems to provide denser feedback during training [3, 6, 7]. Finally, safety remains a paramount concern. Approaches such as Conservative Q-Learning explicitly penalize unsupported actions to reduce distributional shift and avoid unsafe recommendations [3, 8]. Recent work has also investigated the role of temporal discretization in sepsis MDPs, showing that the choice of time-step size can substantially influence policy learning and evaluation outcomes [9].

Despite this progress, several key gaps remain. Nearly all existing studies have focused on adult populations, leaving pediatric sepsis underexplored. Further intensifying the problem, delays in the recognition of the disease, seeking appropriate care, and initiating treatment are common [10] and are especially pronounced in pediatric patients. Early identification of sepsis in children is particularly complicated due to the fact that initial symptoms (such as fever, fast breathing, or irritability) may appear nonspecific [11], leading to worse outcomes when treatment is delayed. Globally, this disease remains a major contributor to morbidity and mortality among children, particularly in critically ill populations [12]. Within pediatric intensive care units (PICUs), sepsis accounts for approximately 8% of admissions and is associated with substantial risk of death and long-term complications. Recent evidence further suggests that mortality among children with sepsis remains considerable even after hospital discharge, with long-term mortality rates estimated at approximately 10% [13, 14]. While this study focuses on in-hospital decision making, these findings highlight the broader burden of pediatric sepsis beyond the acute episode. The time critical nature of the condition, combined with high variability in clinical presentation, underscores the need for improved decision support tools tailored specifically to pediatric care.

The distinction between adult and pediatric sepsis has only recently been formalized. In 2024, the *International Consensus Criteria for Pediatric Sepsis and Septic Shock* introduced a new definition based on the Phoenix Sepsis Score [15]. Under this framework, pediatric sepsis is defined as life-threatening organ dysfunction in the presence of suspected or confirmed infection, quantified by a Phoenix score of at least two points. The score captures dysfunction across four organ systems: respiratory, cardiovascular, coagulation, and neurologic. Septic shock in children is further specified as sepsis accompanied by cardiovascular dysfunction (cardiovascular subscore of at least one point on the Phoenix scale [16]).

For research applications, an extended scoring system known as *Phoenix-8* has been proposed, incorporating four additional organ system domains: endocrine, immunologic, renal, and hepatic. The resulting score enables more precise risk stratification by characterizing the severity of organ dysfunction associated with life threatening illness in pediatric patients. The development of this pediatric specific definition reflects recognition of the unique pathophysiology and clinical presentation of sepsis in children, which differ from those in adults.

Initial attempts to directly transfer the AI Clinician framework architecture to pediatric data have proven challenging [17], highlighting important differences in physiology, disease progression, and treatment protocols. These observations suggest that methods successful in adult settings may not directly generalize to pediatric cohorts without careful adaptation of the underlying modeling choices. In this work, we investigate offline reinforcement learning for pediatric sepsis management using a cohort of 2,229 PICU patients from Great Ormond Street Hospital (GOSH). We focus on two key challenges in clinical RL: reward sparsity and temporal discretization. Specifically, we introduce intermediate rewards derived from the Phoenix-8 scoring system, a marker of mortality risk and illness severity, to provide denser clinical feedback during policy learning. We then compare Double Deep Q-Networks (DDQN) and Conservative Q-Learning (CQL), with and without Phoenix-8–based intermediate rewards, across 4-, 8- and 12-hour temporal resolutions, and evaluate policies through off-policy evaluation and behavioral alignment with physicians. The following methodology follows a structured pipeline, from cohort construction to offline RL policy training and safety evaluation. Figure 1 provides an overview of the full framework employed in this study to train and evaluate different RL models for optimizing sepsis management. Our results highlight temporal discretization as a key lever for stability and safety in offline RL and provide evidence that alternatives beyond the conventional 4-hour setting can improve learning stability while preserving clinically meaningful granularity.

**Fig 1.**
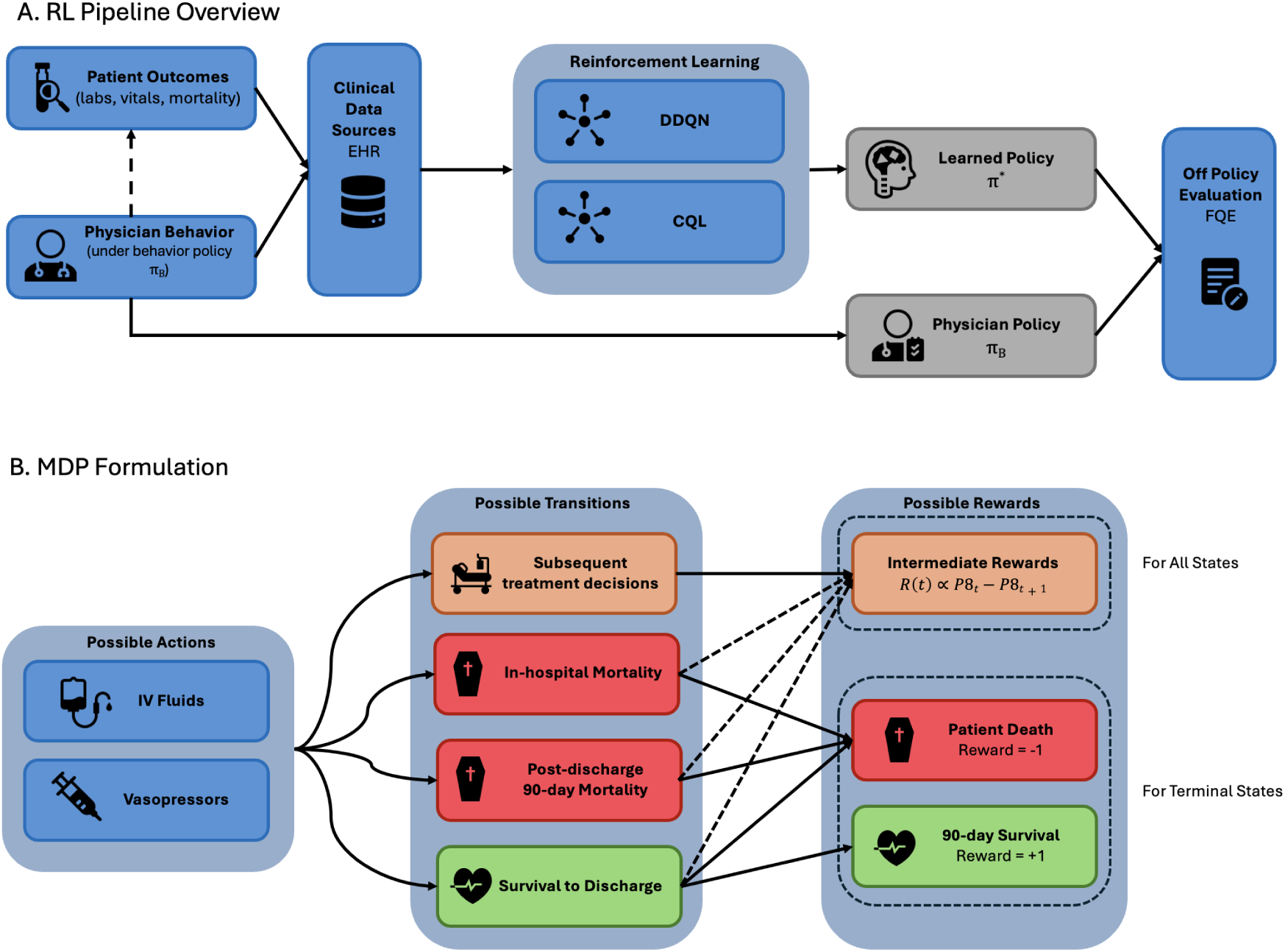
Overview of the reinforcement learning framework. A. The top panel shows the overall RL pipeline from dataset construction to policy evaluation. B. The bottom panel illustrates the Markov Decision Process (MDP) formulation, including actions, transitions and rewards.

## Results

### Cohort Statistics

The GOSH dataset reflects the unique characteristics of a pediatric intensive care unit (PICU) population. The analysis reveals 35.7% of the patients being younger that four months old. This percentage rises to almost half of the cohort when including all patients under the age of one year. The dataset is balanced with respect to sex, with near-equal representation of male and female patients, ensuring that model training is not biased by sex specific imbalances. Within 90 days of ICU admission, 9.6% of patients died, including both in-hospital and post-discharge mortality. This mortality rate is lower compared to that of adult sepsis cohorts reported across studies [4].

The final cohort used in this study is summarized in Table 1. The table presents key demographic and clinical variables stratified by survival status, allowing comparison between patients who survived and those who did not. Non-survivors were younger on average (3.6 vs. 4.1 years) and included a smaller proportion of female patients (49.3% vs. 57.1%). Non-survivors also exhibited higher Phoenix-8 scores at both the start (2.73 vs. 2.17) and end of their series (4.48 vs. 3.54), indicating a greater degree of organ dysfunction. Additional differences were observed in urine output, which was lower in non-survivors.

**Table 1.** Cohort characteristics for the final dataset stratified by survival status. Values are presented as means unless otherwise indicated.

| Variable | Survival (n = 2016) | Non-survival (n = 213) |
| --- | --- | --- |
| Female (%) | 57.1 | 49.3 |
| Mean age (years) | 4.1 | 3.6 |
| Mean weight (kg) | 15.67 | 13.06 |
| Phoenix-8 score at series start (min / avg / max) | 0 / 2.17 / 10 | 0 / 2.73 / 11 |
| Phoenix-8 score at series end (min / avg / max) | 0 / 3.54 / 12 | 0 / 4.48 / 11 |
| Platelet count ($\times 10^3/\mu\text{L}$) | 219.3 | 196.0 |
| Total bilirubin ($\mu\text{mol/L}$) | 31.9 | 42.2 |
| Mean blood pressure (mmHg) | 61.9 | 60.6 |
| Glasgow Coma Scale (points) | 11.4 | 11.1 |
| Creatinine ($\mu\text{mol/L}$) | 72.6 | 50.3 |
| Urine output (ml/4h) | 64.2 | 59.0 |

### CQL Learns Safer Sepsis Treatment Policies

In line with prior work on sepsis RL, we first evaluate all policies using the conventional 4-hour temporal resolution. CQL demonstrated superior stability and convergence across all metrics. While DDQN often suffered from exploding Q-values and collapsing discounted returns (indicating a failure to generalize from offline data) CQL maintained controlled growth and stable performance.

Behaviorally, Figure 2 shows CQL policies aligned closely with clinician practice. For vasopressor decisions, CQL achieved high action-level similarity (78-79% agreement with physician decisions), and for fluids it reproduced the cautious dosing patterns observed in the dataset (83–86% agreement), avoiding extreme over-resuscitation. DDQN policies, by comparison, diverged more frequently from clinician actions (similarity never exceeded 15–24%), sometimes favoring aggressive fluid strategies or atypical action combinations, particularly in severe or rapidly changing states. Similar patterns were observed at 8-hour and 12-hour temporal resolutions (see Supplementary Figure S1).

**Fig 2.**
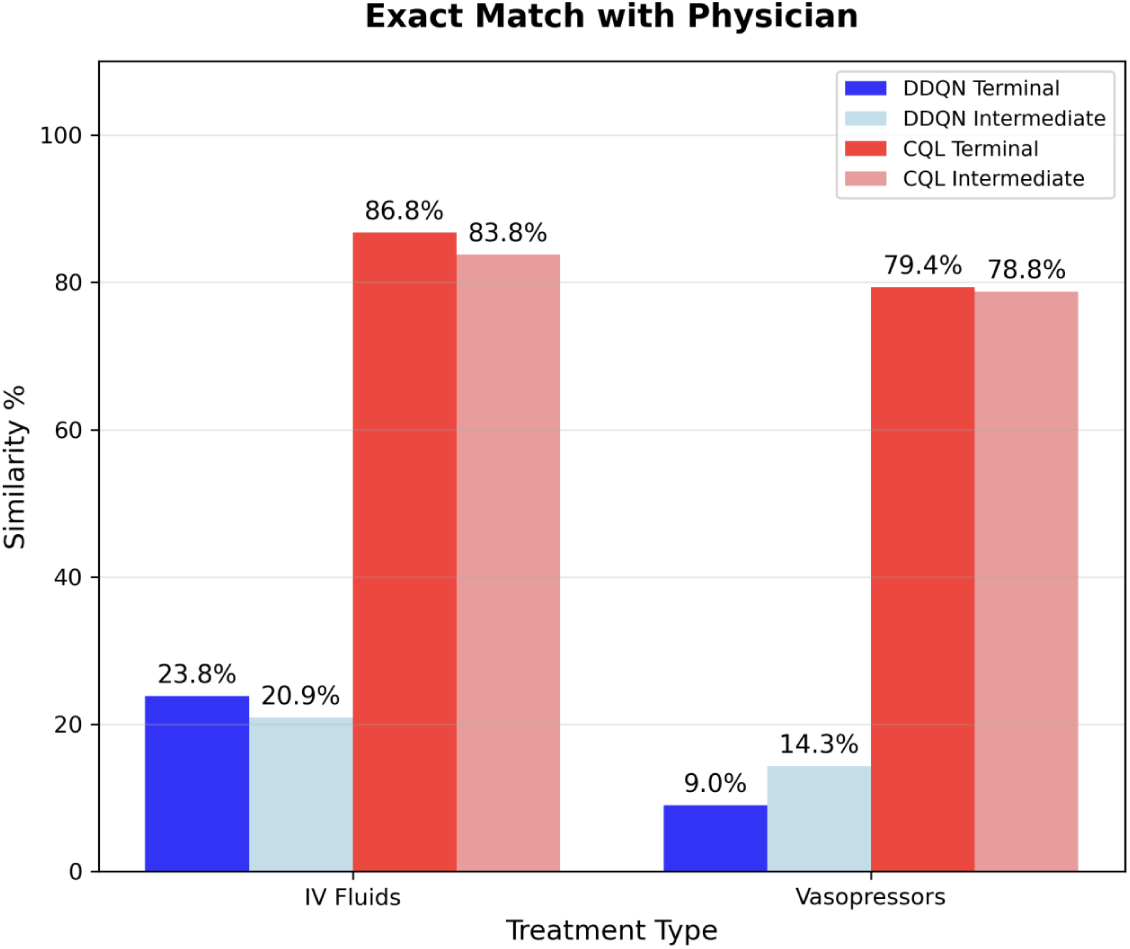
Action similarity between RL policies and physician decisions at 4-hour temporal resolution. CQL (red) demonstrates substantially higher agreement with clinician vasopressor and fluid decisions compared to DDQN (blue). Light shades represent intermediate reward variants, while bright shades represent terminal reward variants.

Severity stratified treatment trajectories (Figure 3) show that CQL scales interventions with illness severity, using minimal vasopressor support in mild cases and escalating treatment intensity at higher Phoenix-8 scores. In contrast, DDQN exhibits weaker discrimination across severity strata and more erratic dosing patterns, evidenced by data in Figure 3. When stratifying by outcome, CQL trajectories for patients who ultimately died remain closely aligned with historical clinician behavior, suggesting that conservative offline RL in this cohort largely reproduces standard care rather than proposing substantially different treatment strategies.

**Fig 3.**
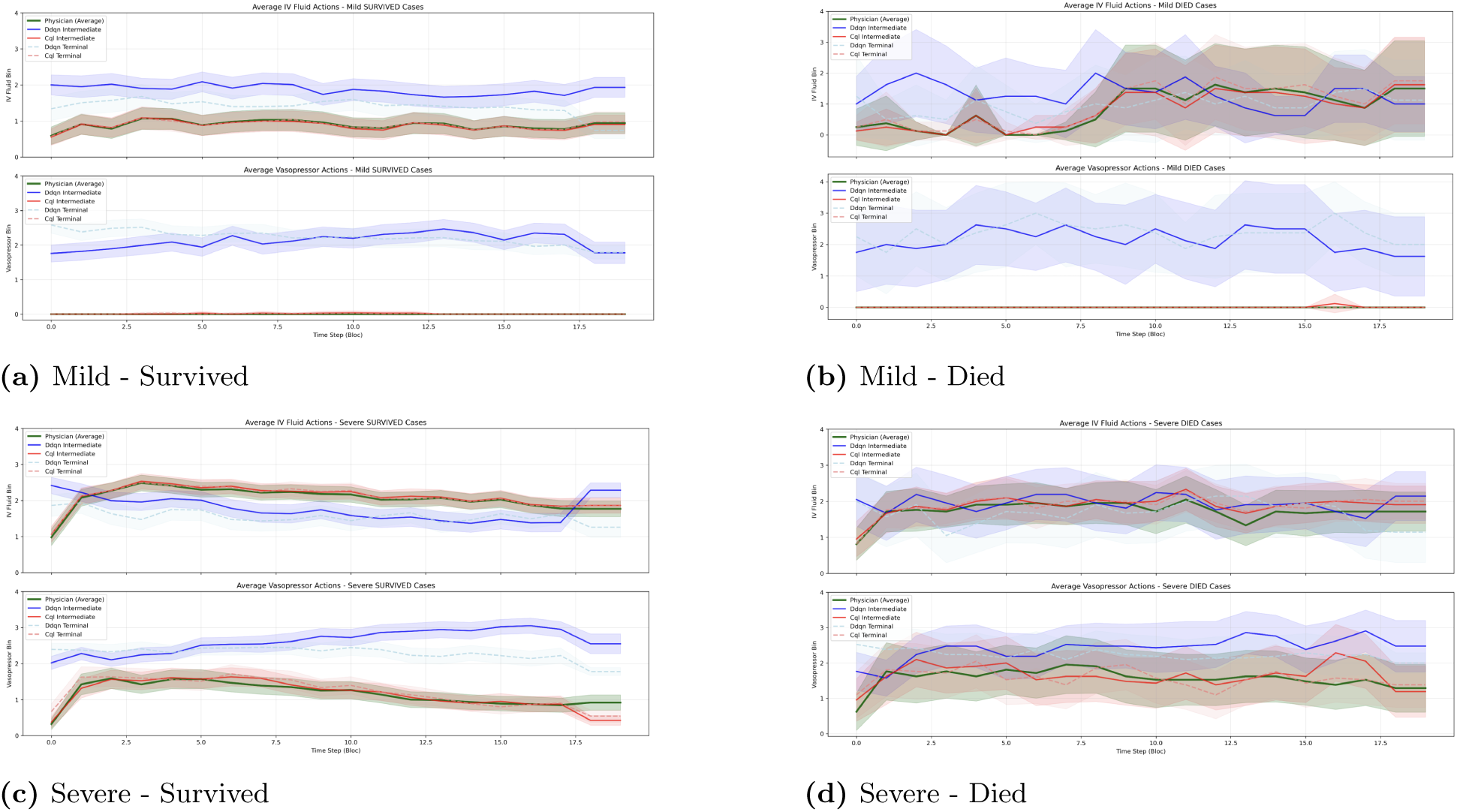
Average treatment decisions for vasopressors and intravenous fluids, comparing RL policies and physician actions (shown in green) across patient severity and outcomes. Mild cases are presented in the top row and severe cases in the bottom row; survived patients are in the left column, deceased patients in the right column. CQL models (red tones) generally adjust interventions in line with patient severity, increasing vasopressor use for severe cases and reducing interventions for mild cases. DDQN models (blue tones) show less discrimination by severity. For patients who ultimately died, CQL policies largely mirror physician actions, indicating that the models do not deviate from standard practice even in adverse outcomes. These trajectories correspond to 4-hour binning; similar patterns were observed at other temporal resolutions.

Action-distribution plots at 4-hour resolution (Figure 4) further highlight these differences: DDQN tends to favour combinations of low vasopressor use with relatively high fluid doses, whereas CQL maintains an action distribution that closely matches physician practice and becomes more conservative in its fluid recommendations.

**Fig 4.**
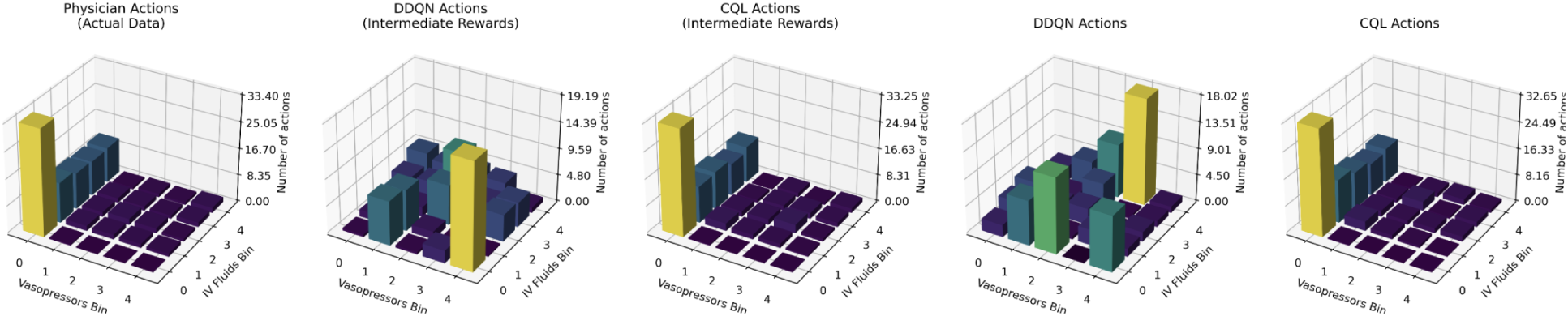
Three-dimensional distributions of RL policy actions for vasopressor and intravenous fluid administration.

Despite these favourable results for CQL at 4-hour resolution, the choice of reward function plays a critical role in shaping policy behaviour. We observed that intermediate rewards based on changes in Phoenix-8 did not consistently align with terminal outcomes. As shown in Figure 5, the mean trajectories of severe patients who survived demonstrate an initial worsening in Phoenix-8 scores (resulting in negative intermediate rewards) followed by subsequent stabilization and improvement. This pattern was consistently observed across survivor groups and likely reflects the clinical reality that early therapeutic interventions can transiently exacerbate physiological indicators before yielding longer-term benefit.

**Fig 5.**
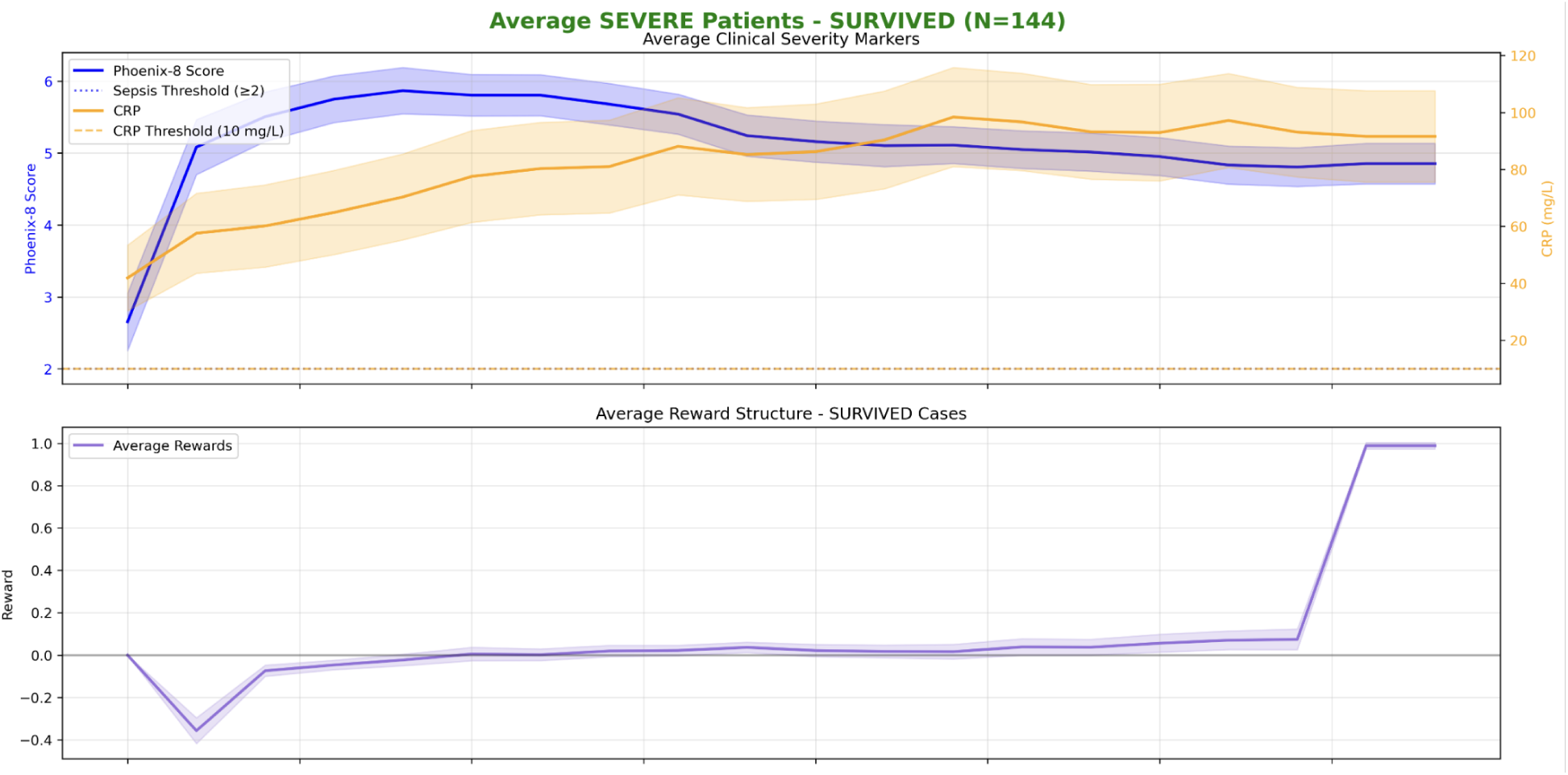
Average trajectories for Phoenix-8 score, C-reactive protein (CRP), and intermediate rewards in severe patients who survived. The trajectories show an initial increase in organ dysfunction and inflammatory markers, corresponding to negative intermediate rewards, followed by stabilization or improvement over time.

Phoenix-8 remained relatively stable across consecutive 4-hour timesteps, largely due to the infrequent updating of its constituent variables (particularly laboratory measurements). As a result, reward signals were often unchanged between transitions reducing the effectiveness of intermediate reward shaping and diminishing the distinction between intermediate and terminal reward formulations. While intermediate rewards did not consistently alter training dynamics at the 4-hour resolution, they occasionally led to higher-performing policies in relative terms. Notably, CQL with intermediate rewards achieved the strongest performance compared to the behaviour policy at this resolution. However, this improvement appears sensitive to noise in the reward signal, as Phoenix-8 changes remain sparse and temporally misaligned with clinical improvement. As such, intermediate rewards at 4-hour bins may provide opportunistic gains rather than a consistently reliable learning signal.

These findings suggest that the utility of intermediate rewards is closely linked to the temporal discretization of the data. We therefore examined whether altering the temporal resolution can improve learning stability and policy behaviour.

### Temporal Aggregation Stabilizes Offline RL

Temporal binning emerged as a major factor in model success. Moving from 4-hour to 8- and 12-hour bins shortened episode horizons and reduced the number of decision points per trajectory, as summarized in Table 2. Larger sampling intervals produced fewer time steps and shorter average episodes, effectively simplifying the decision process while preserving the same underlying clinical window.

**Table 2.**
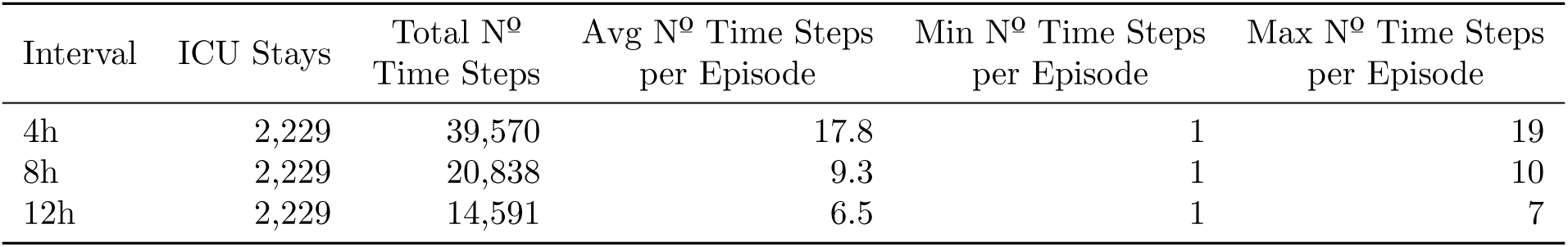
Episode length summary across time binning intervals. Values represent averages over all patient episodes.

To quantify how time-step size affected rewards, we examined reward statistics across temporal resolutions. Coarser binning reduced the variance of total intermediate returns across patients (from 0.4902 at 4h to 0.4842 at 8h and 0.4805 at 12h), indicating a cross-patient smoothing effect, while terminal reward variance remained constant at 0.3450, as expected given unchanged outcomes. In contrast, within-patient variability increased with coarser bins: the average standard deviation of intermediate rewards rose from 0.262 to 0.359 and 0.428 for 4h, 8h, and 12h respectively, and from 0.233 to 0.316 and 0.371 for terminal rewards. These patterns suggest that temporal aggregation smooths trajectories between patients but makes individual reward jumps larger as samples move further apart in time.

At the algorithmic level, these changes in horizon and reward structure translated into different learning dynamics (Figure 6). For CQL, increasing the bin size produced visibly more stable training curves, with smoother temporal-difference losses and more compact Q-value distributions, particularly at 8-hour resolution. For DDQN, coarser binning mitigated but did not eliminate instabilities associated with extrapolation: Q-values remained prone to overestimation and oscillations, especially when combined with intermediate rewards.

**Fig 6.**
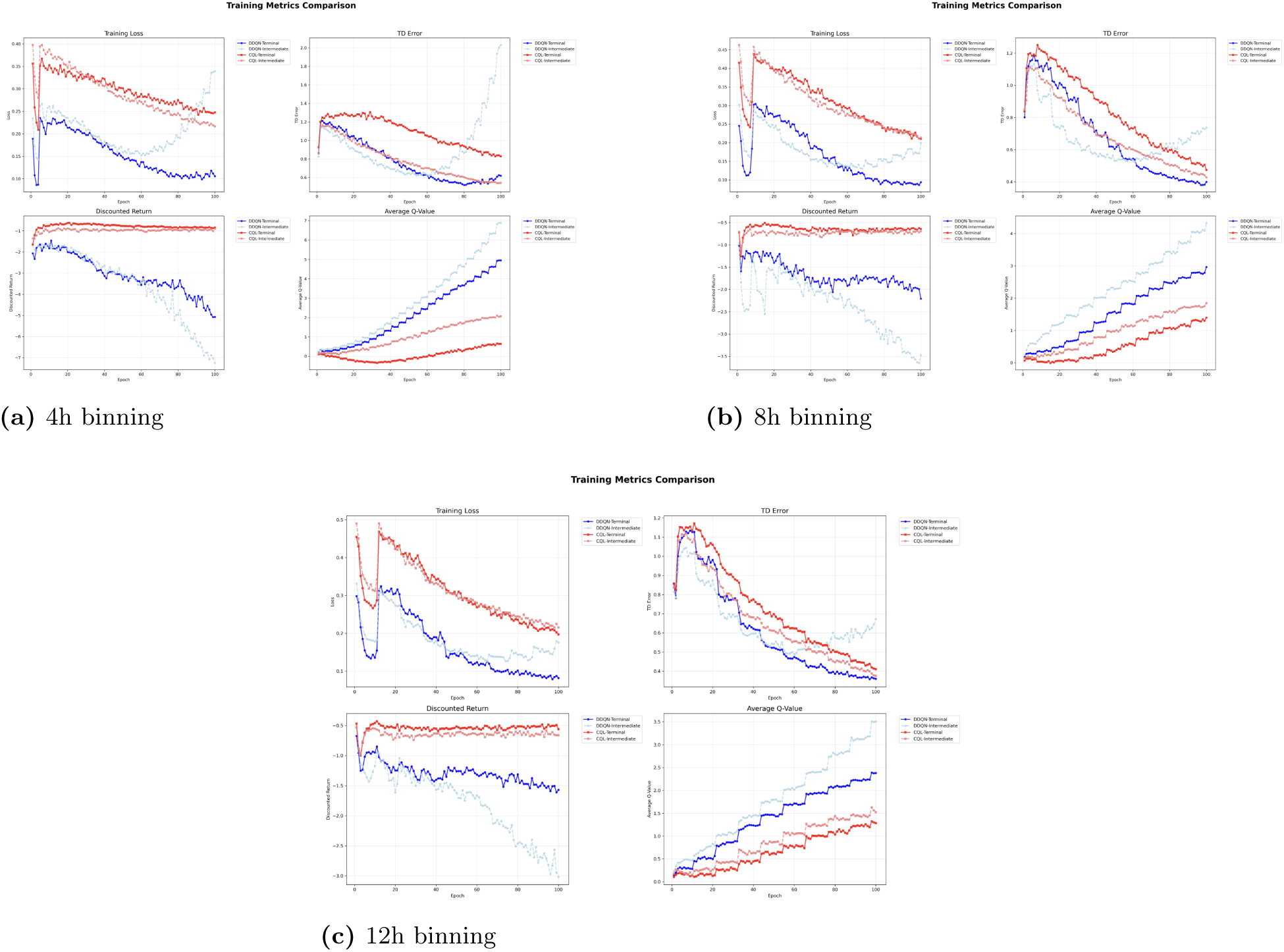
Training performance of all reinforcement learning models at different temporal resolutions: (a) 4-hour, (b) 8-hour, and (c) 12-hour. Each subplot depicts the evolution of training loss, TD error, estimated Q-values, and discounted returns across epochs. DDQN models, in blue, demonstrates marked instability with spiking losses and TD error, and exploding Q-values. CQL variants, in red, exhibit stable and conservative convergence, with decreasing losses, controlled Q-value growth, and stable discounted returns. 4-hour bins worsened DDQN instability, while increasing bin size smoothed the trajectories and converges to lower loss and TD error values.

At the policy level, 8-hour binning emerged as the most favourable compromise. This selection is supported not only by absolute performance but by relative improvement over the behaviour policy. Training stability and performance were evaluated through Fitted Q Evaluation (FQE) convergence plots (see Supplementary Figure S2). At 8-hour resolution, CQL (without intermediate rewards) demonstrated a substantially larger gain over behaviour cloning compared to 4-hour bins (this pattern is reflected in FQE estimates in Fig 7), indicating that the learned policy is not merely reproducing clinician actions but extracting additional value from the data.

**Fig 7.**
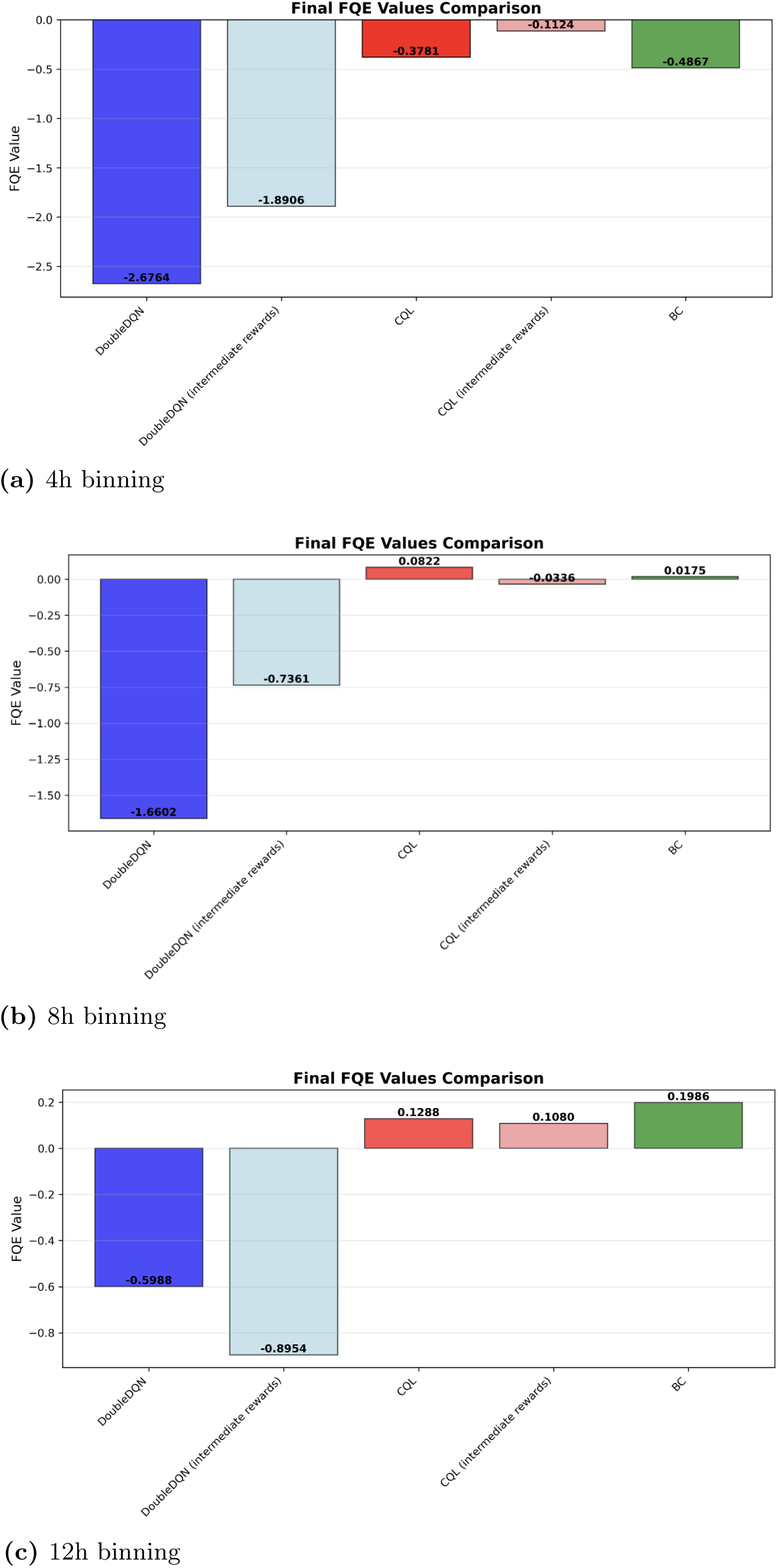
Fitted Q-Evaluation (FQE) estimates across different temporal resolutions: (a) 4-hour, (b) 8-hour, and (c) 12-hour. Values (approximately bounded between −1 for death and +1 for survival) indicate expected returns, with higher values reflecting better outcomes. Results are not directly comparable across resolutions (*γ* = 0.99). Across all resolutions, BC (green) and CQL (red tones) achieved the highest FQE values, while DDQN variants (blue tones) consistently ranked lowest.

In contrast, improvements at 4-hour resolution were smaller and more sensitive to reward specification. While finer temporal granularity preserves detail, it also amplifies noise in Phoenix-8 changes and lengthens horizons. This increases sensitivity in value estimations particularly for non-conservative methods such as DDQN.

At 12-hour resolution, further aggregation stabilized training but increased the magnitude of within-patient reward transitions, effectively compressing multiple clinically relevant decisions into single steps. This risks obscuring meaningful temporal structure and limiting the ability of the model to associate interventions with outcomes.

Taken together, these results support viewing time-step size as a primary design choice in offline RL for healthcare rather than a purely technical detail inherited from prior work. Coarser, clinically motivated temporal aggregation can act as a regularizer that improves stability and safety, but overly aggressive aggregation may discard information needed for nuanced decision making, underscoring the need to tune temporal resolution jointly with algorithm and reward design.

## Discussion

A primary takeaway of this study is that conservative offline reinforcement learning algorithms like CQL provide significantly greater stability and reliability than standard deep RL approaches when applied to real world pediatric ICU data. In our experiments, while DDQN frequently exhibited unstable training dynamics (including exploding Q-values and highly variable discounted returns), CQL maintained controlled value estimates and stable convergence across temporal resolutions. Beyond algorithmic stability, CQL policies also demonstrated substantially greater behavioral alignment with physician decisions than DDQN policies. This alignment is particularly important in high stakes clinical settings, where deviations from established practice can lead to unsafe or implausible recommendations. By explicitly penalizing out-of-distribution actions, CQL ensures that recommendations stay within the safe zone of established medical practice.

However, the behaviour of these models is fundamentally shaped by the design of the reward function. In clinical reinforcement learning, rewards serve as proxies for patient improvement, yet defining appropriate reward signals remains challenging. The need for intermediate rewards is often motivated by the sparsity of terminal outcomes, such as survival, which provide limited and delayed feedback. As discussed by Brian Christian in his book “The Alignment Problem” [18], sparse reward structures can force models to explore unsafe or irrelevant behaviours before receiving meaningful feedback. In this work, we use Phoenix-8 as an intermediate reward to provide structured feedback on organ dysfunction. Phoenix-8 captures clinically relevant physiological domains and offers a meaningful proxy for patient status. However, its components are not uniformly or continuously observed. While some variables, such as cardiovascular and respiratory measures, are frequently monitored, others, particularly laboratory values, are collected intermittently and at the discretion of the clinical team. This introduces temporal sparsity in the reward signal and couples the reward itself to clinician driven measurement patterns.

The relationship between intermediate rewards and terminal outcomes reflects a broader challenge in clinical reinforcement learning. Improvements in Phoenix-8 are intended to represent short-term recovery in organ dysfunction, yet such improvements do not always correspond to better long-term outcomes such as survival. In some cases, interventions that transiently improve physiological markers may not lead to improved patient trajectories. This highlights the intrinsic trade-off in reward design: terminal outcomes provide clinically meaningful but sparse signals, while intermediate rewards offer denser feedback but may emphasize short-term physiological targets. Based on our findings, policies trained with either intermediate or terminal rewards produced broadly similar treatment patterns suggesting that both formulations capture related aspects of clinical decision making. However, the limited variability of Phoenix-8 at the 4-hour resolution further constrained the effectiveness of intermediate rewards. As a result, the reward signal often remained unchanged across consecutive timesteps, reducing its ability to meaningfully shape policy learning.

Taken together, these findings indicate that the effectiveness of intermediate rewards is closely tied to how frequently they are observed and updated. This observation motivates the consideration of temporal discretization as a key design choice in clinical reinforcement learning. In our setting adjusting the temporal resolution directly impacts both the informativeness of the reward signal and the stability of learning. Coarser temporal aggregation may act as a form of regularization smoothing noisy trajectories and aligning decision points with clinically meaningful updates, while finer resolutions risk amplifying noise and reducing reward informativeness. In contrast to prior work on dense adult ICU datasets, where finer time resolutions may be advantageous [9], our results suggest that pediatric datasets with sparser observations benefit from longer time intervals between decision steps.

The potential role of artificial intelligence in sepsis management remains both promising and controversial. On one hand, RL methods offer a framework for learning personalized treatment policies from large clinical datasets and may eventually help clinicians navigate complex therapeutic trade-offs. On the other hand, concerns about safety and interpretability remain significant. Models such as DDQN can generate extreme or clinically unrealistic treatment suggestions, particularly when exposed to noisy or incomplete data. Even conservative approaches such as CQL primarily reproduce existing clinical behavior rather than discovering fundamentally new treatment strategies. In this sense, the models developed here function largely as physician imitators rather than innovators. However, in high stakes clinical domains, establishing reliable and clinically aligned decision support systems is a necessary first step before more exploratory policy learning can be considered.

Pediatric sepsis remains one of the most challenging conditions in critical care medicine, characterized by rapid progression, high mortality risk, and the need for immediate, precise clinical decision making. Traditional approaches to sepsis management rely heavily on standardized guidelines, clinician heuristics, and scoring systems that may not capture the full complexity of individual patient trajectories. The heterogeneous nature of pediatric populations, combined with the time critical nature of sepsis intervention, creates an urgent need for data-driven approaches that can support clinicians in making optimal treatment decisions.

The findings of this study must be interpreted within the context of several limitations inherent to the dataset and methodological design. First, GOSH is a highly specialized pediatric hospital without an emergency department and many patients are referred from other institutions for complex or severe conditions, a demographic that may not be representative of other PICU populations globally. This poses a challenge for external validity, as the model’s learned policy may be biased towards specific physiological responses observed at GOSH. Second, the low mortality rate, while clinically positive, may prejudice the reinforcement learning agent, as it provides a sparse and infrequent signal for learning about negative outcomes. Third, although CQL improves safety by restricting policies to actions supported by the dataset, this conservative bias may also limit the discovery of genuinely superior treatment strategies. In contrast, exploratory algorithms like DDQN are more capable of identifying novel policies but are also substantially more vulnerable to instability and noise.

## Materials and Methods

### Ethics Statement

This retrospective study used de-identified patient data accessed through the Great Ormond Street Hospital (GOSH) Data Research Environment (DRE). Ethical approval for the underlying project was provided by the National Health Service (NHS) Health Research Authority (HRA) (IRAS ID: 301663, REC reference 21/LO/0646). The authors had access only to de-identified data and did not have access to information that could identify individual participants during or after data collection. Data were accessed for the purposes of this study from 15/05/2024 to 08/09/2025.

### Dataset and Cohort Construction

The construction of the data set and the definition of the cohort follow previous work on this cohort [17]. The initial data extraction was restricted to patient IDs associated with a historical hospital diagnosis containing the keyword “sepsis”. This filtering step was necessary due to the large size of the underlying dataset (approximately 14 million records) and computational constraints that made it impractical to process the full hospital population spanning 2000–2024. However, diagnosis labels alone were insufficient to define sepsis episodes, as they are linked to patient identifiers rather than specific ICU stays, and historical diagnoses may not align with contemporary sepsis definitions.

To address this, sepsis episodes were defined using the Phoenix-8 scoring system applied directly to ICU physiological data. Specifically, ICU stays were retained if the Phoenix-8 score exceeded 2, indicating organ dysfunction consistent with the clinical definition of sepsis. The Phoenix-8 scoring system was designed for pediatric populations and integrates multiple physiological variables to quantify the severity of organ dysfunction.

CRP was incorporated as an additional filter to capture episodes associated with significant inflammatory responses. However, CRP is a non-specific biomarker of inflammation and may be elevated in a variety of clinical contexts, including sterile inflammation such as the postoperative state. Consequently, the CRP threshold should be interpreted as a pragmatic filtering criterion rather than definitive evidence of infection. After applying the Phoenix-8 ≥ 2 and CRP ≥ 10 mg/L criteria [17], 2,229 unique ICU stays were retained for analysis.

For each episode, we extracted a 72-hour window spanning up to 48 hours before and 24 hours after sepsis onset. However, only 17.45% of patients had complete information for the 48 hours preceding onset, a limitation largely explained by the fact that many patients at GOSH are admitted as external referrals [17]. To mitigate excessive data loss, we adopted a flexible windowing strategy anchored at sepsis onset: for each patient, the window extends backwards and forwards from onset to include up to 72 hours of available data, prioritizing pre-onset observations when present, and otherwise extending further into the post-onset period when pre-onset data are limited. As a result, trajectories may contain varying proportions of pre- and post-onset data across patients.

While this approach increases cohort retention, it introduces heterogeneity in temporal coverage which may influence downstream learning by exposing models to different phases of disease progression across trajectories. Nevertheless, for a specialized cohort like GOSH (where many patients have a complex medical history and may be recovering from surgery) high baseline levels of markers like CRP or even the Phoenix-8 score may not be directly indicative of sepsis alone. Restricting trajectories to a focused 72-hour window therefore prioritizes the acute phase of sepsis while avoiding confounding from longer-term disease processes. Although this truncation reduces the overall number of time steps and may not fully capture recovery dynamics, it represents a clinically grounded compromise between data completeness and relevance.

### MDP Formulation

Sepsis provides a compelling case study for reinforcement learning because clinical treatment naturally unfolds as a sequence of decisions under uncertainty. At each point in time, a clinician observes a patient’s physiological state (vital signs, laboratory results, organ function) and must decide on an appropriate intervention, such as administering fluids, adjusting vasopressors, or giving antibiotics. These treatment decisions correspond to the actions in a MDP, while the evolving patient state defines the environment.

#### State Space

The dataset was preprocessed to include 47 features encompassing demographics, vital signs, and lab results (see Table S1 for the complete list). These features were derived from 157 raw fields with documented outlier removal and imputation using sample-and-hold, interpolation, k-nearest neighbors, and explicit missing-value indicators; the feature consolidation, outlier criteria (Table S2), propagation windows (Table S3), and missing-value handling strategies follow prior work on this cohort [17].

#### Action Space

The action space captured joint intravenous fluid and vasopressor decisions, with vasopressor doses aggregated into a Vasoactive Inotropic Score (Equation S1) aligned with GOSH practice and mapped, together with fluid volume, onto a 5×5 discretized grid of 25 actions (Table S4). We excluded vasoactive agents not used routinely at GOSH and piggyback fluid medications, focusing on the main resuscitation levers of PICU sepsis care [17].

#### Reward Signal

The use of Phoenix-8 as an intermediate reward signal in pediatric sepsis was proposed and statistically validated in prior work on this cohort [17], with the specific functional form adapted from a previously developed reward mechanism for adults with sepsis [3]. Rewards combined terminal 90-day outcomes with optional Phoenix-8-based intermediate shaping. Terminal rewards were +1 for survival and −1 for death, while intermediate rewards scaled changes in Phoenix-8 between consecutive time steps, rewarding improvement and penalizing deterioration, with normalization by cohort-level Phoenix-8 range and a Heaviside term to emphasize improvements. The adapted reward formulation is given by:

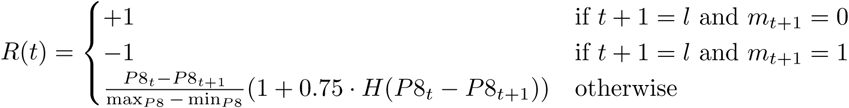

where *P* 8*_t_* is the Phoenix-8 score at timestep *t*, *m_t_* equals 1 if the patient has died and 0 otherwise, *l* denotes the ICU episode length, max*_P_* _8_ and min*_P_* _8_ are the observed bounds of Phoenix-8, and *H* is the Heaviside step function.

This design allows the agent to receive positive reinforcement not only at the terminal outcome (survival) but also for intermediate improvements in clinical severity. In contrast, the severity of worsening is penalized. Given the rapid progression of sepsis, these intermediate signals are crucial for shaping learning and guiding the agent toward clinically consistent decision making while preserving the long-term focus on survival.

### Algorithm Selection

We considered two offline value-based methods: Double Deep Q-Networks (DDQN) and Conservative Q-Learning (CQL). DDQN is value-based method that decouples action selection from evaluation to reduce overestimation bias, while CQL is a conservative variant that penalizes Q-values for actions not present in the dataset, mitigating the risk of distributional shift and unsafe recommendations.

#### Double Deep Q-Network (DDQN)

DDQN was chosen as a natural extension of Deep Q-Networks (DQN), one of the foundational algorithms in reinforcement learning. DQN integrates Q-learning with deep neural networks to approximate the action–value function, enabling the handling of high-dimensional state spaces that are common in healthcare settings with rich patient data [19].

Formally, the DDQN update is given by Equation 1, where the next action is chosen according to the current Q function but evaluated using the target network [20]:

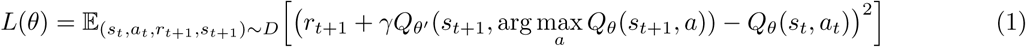

#### Conservative Q-Learning (CQL)

In contrast to DDQN, which can extrapolate optimistically beyond observed behavior, CQL is explicitly designed to mitigate such risks. The update formula resembles that of DQN but includes an additional regularization term that penalizes large Q-values for actions that are not well supported by the dataset. Specifically, CQL regularizes, in expectation, the magnitude of Q-values for out-of-distribution (OOD) actions by pushing them downward relative to those observed in the data [21]. The degree of this conservatism is controlled by the parameter *α* where larger values push the learned policy closer to the behavior policy [22].

In this study, the discrete version of CQL (Equation 2) was adopted because the action space corresponds to a finite set of clinical treatment options rather than continuous dose ranges. Each decision step involves categorical choices such as administering intravenous fluids, initiating vasopressors, or withholding intervention, which aligns naturally with the discrete formulation.

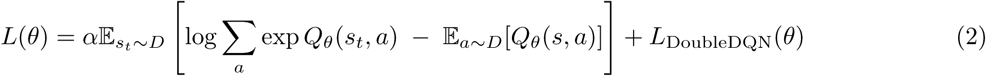

CQL has shown strong performance in sepsis treatment tasks [3, 8], where safety is essential. By introducing a penalty term into the loss function, CQL balances performance with caution, promoting clinically plausible recommendations and reducing the likelihood of unsafe out-of-distribution actions.

### Experimental Setup

#### Data Splitting

The dataset was divided into training and testing subsets using an 80/20 stratified split. Stratification was performed across two clinically relevant axes, mortality outcome and episode length, ensuring balanced representation and preserving distributional consistency between splits. This is especially important in clinical datasets, where outcome imbalances are common and may bias evaluation.

#### Model Variants

This design resulted in four main policy variants: *DDQN-terminal, DDQN-intermediate, CQL-terminal, and CQL-intermediate*. All models were implemented using the d3rlpy library. The selected hyperparameters were chosen based on prior literature in medical RL, standard practices in deep reinforcement learning, and preliminary experiments on the dataset. To ensure comparability across algorithms, hyperparameters were kept fixed across model variants, and training was monitored to confirm that all networks converged according to their respective training metrics.

Model checkpoints were selected based on the stabilization of the training curves. Specifically, the epoch at which performance plateaued without further improvement was selected as the final model checkpoint, reducing the risk of selecting undertrained or unstable policies. Table 3 summarizes the key settings used in training.

**Table 3.** Hyperparameter configuration used for all RL models.

| Hyperparameter | Value |
| --- | --- |
| Learning rate | $1 \times 10^{-4}$ |
| Discount factor ($\gamma$) | 0.99 |
| Batch size | 128 |
| Training epochs | 100 (policy), 200 (FQE) |
| Gradient clipping | L2 norm at 1.0 |
| Target network update interval | 1000 steps |
| Steps per epoch | $\lceil \frac{n_{\text{obs}}}{\text{batch size}} \rceil$ |
| Total steps | Steps per epoch × #epochs |

#### Baseline Model

A Behavioral Cloning (BC) policy was trained to approximate clinician decisions and serve as a lower-bound reference for evaluation. While BC is limited to imitation and cannot exceed observed clinical performance, it provides a valuable “standard of care” comparator when assessing the potential benefits and risks of RL-derived policies.

#### Training Monitoring

Model training and convergence were tracked across multiple metrics:

- **Training loss:** standard optimization progress indicator.

**–** For DDQN this corresponds to the squared TD error.
**–** For CQL the loss additionally includes the conservative regularization term that penalizes high Q-values for out-of-distribution actions.
- **Temporal Difference (TD) error:** reduction indicates improved value function approximation.
- **Discounted return:** estimates of expected cumulative reward.
- **Average Q-values:** stabilization indicates convergence.

Together, this setup ensured that algorithmic performance could be compared fairly across methods and reward designs, while grounding evaluations in clinically meaningful baselines.

### Policy Evaluation & Safety

#### Off-Policy Evaluation (OPE)

We evaluated the performance of the learned policy using OPE, specifically Fitted Q Evaluation (FQE). FQE works by training a separate value function to approximate the expected returns of a given policy, using the available offline data. By doing so, it provides a principled way to compare candidate policies, even when their recommendations differ from those observed in practice. FQE is particularly well suited for healthcare applications because it has demonstrated superior performance in accurately ranking policies and robustness across challenging scenarios [23]. This ensured that the policies could be quantitatively evaluated while preserving patient safety. FQE was trained on the test dataset using the same hyperparameter configuration as policy training, but with extended training duration (200 epochs) to improve value function estimation and ensure model convergence. The evaluation computed Q-values for initial state–action pairs from test episodes, yielding policy value estimates that enabled direct comparison across model variants.

#### Behaviour Comparison

To assess clinical plausibility, model action distributions were systematically compared against physician practice patterns. The primary metrics were the proportion of time steps where the policy exactly matched the clinician’s action and the overall frequency of each action across the 25-action space. Beyond aggregate statistics, patient trajectories were stratified by severity (mild: Phoenix-8 ≤ 3, moderate: 4–5, severe: ≥ 6) and visualized to analyze Phoenix-8 score evolution, treatment decisions, and areas of agreement or disagreement between model and physician actions. This allowed both quantitative and qualitative assessment of how closely RL policies aligned with standard clinical practice.

## Acknowledgments

This research was funded by GOSH Children’s Charity Award VS0618.

## Data Availability Statement

The data that support the findings of this study are not publicly available due to patient confidentiality and institutional restrictions. The data are stored within the Great Ormond Street Hospital (GOSH) Data Research Environment (DRE) and may be made available to qualified researchers on reasonable request, subject to appropriate approvals and data sharing agreements.

## Author Contributions

RW designed and implemented the data extraction and processing pipeline (including cohort definition based on the Phoenix-8 and CRP criteria), designed and implemented the state and action representations, proposed and statistically validated the use of the Phoenix-8 score as an intermediate reward signal, and conducted preliminary experiments with the AI Clinician approach that informed model selection. FBM conceptualized the study, designed the reinforcement learning approach in collaboration with PM and with input from JS, implemented the reinforcement learning pipeline, conducted the evaluation analyses, and drafted the manuscript. PM supervised the study and provided guidance on artificial intelligence methodology. JS contributed with expertise and review on reinforcement learning methods. SR provided clinical expertise in pediatric sepsis and contributed to the adoption of the Phoenix-8 criteria. MP provided clinical insight as a professor of pediatric intensive care. All authors reviewed and approved the final manuscript.

## A Supplementary Notes

### Vasoactive Inotropic Score

The Vasoactive Inotropic Score (VIS) was used to aggregate vasopressor doses into a universal feature, following the pediatric-specific construction developed in prior work [17]. The formula for VIS is provided below, with a factor of 2.5 applied to the vasopressin component to account for unit conversion.

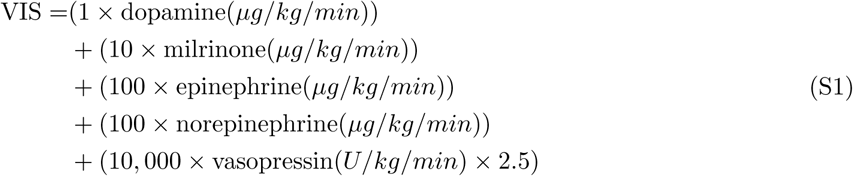

## B Supplementary Tables

**Table S1.**
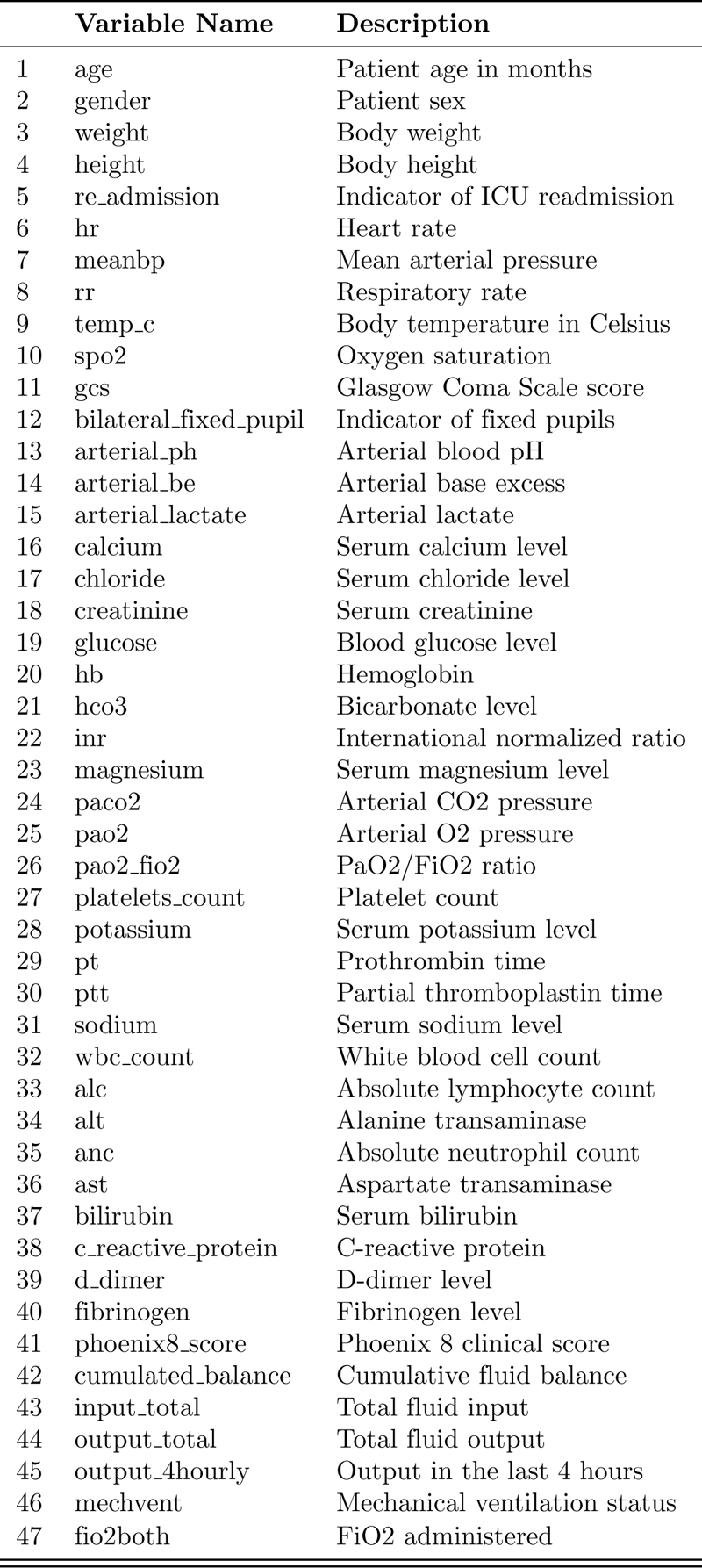
Variables used in the analysis, following prior work on this cohort [17].

**Table S2.** Outlier criteria applied to the dataset [17].

|  | Variable Name | Criterion Type |
| --- | --- | --- |
| 1 | Weight | > 200 (kg) |
| 2 | Height | > 200 (cm) |
| 3 | Mean Arterial Pressure (MAP) | > 200 (mmHg) |
| 4 | Glasgow Coma Scale (GCS) | < 3 or > 15 |
| 5 | Respiration Rate | > 200 (breaths per minute) |
| 6 | Heart Rate | > 600 (beats per minute) |
| 7 | Temperature | < 22 or > 47 (°C) |
| 8 | Lactate | > 50 (mmol/L) |
| 9 | Creatinine | > 2000 ($\mu\text{mol/L}$) |
| 10 | Bilirubin | > 850 ($\mu\text{mol/L}$) |
| 11 | Sodium | < 95 or > 250 ($\mu\text{mol/L}$) |
| 12 | Potassium | < 1.5 or > 10 (mmol/L) |
| 13 | Arterial pH | < 6 or > 8.5 |
| 14 | PaO2 | < 1 or > 100 (mmHg) |
| 15 | FiO2 | < 21 or > 100 (% oxygen) |
| 16 | SpO2 | < 0 or > 100 (% oxygen saturation) |

**Table S3.** Propagation windows for different clinical feature groups. The feature-family-specific propagation strategy and window lengths follow prior work on this cohort [17], where each window is justified on the basis of the clinical relevance and measurement frequency of the feature group.

| Feature Group | Variables | Window |
| --- | --- | --- |
| Static Features | Weight, Height | Max-out (entire ICU stay) |
| Metabolic Indicators | Lactate, HCO<sub>3</sub>, Arterial pH | 8 hours |
| Electrolytes | Glucose, Chloride, Sodium | 16 hours |
| Coagulation Markers | Platelets, INR, D-dimer, Fibrinogen, PT, PTT | 28 hours |
| Renal, Hepatic, Immune Markers | Creatinine, Bilirubin, ALT, AST, CRP, ANC, ALC, Magnesium, Calcium | 28 hours |
| Long-term Electrolyte Balance | Potassium | 144 hours |
| Blood Cell Stability | Hemoglobin (Hb), White Blood Cells (WBC) | 28 hours |
| Platelet Dynamics | Platelets | 16 hours |
| Vital Signs | MAP, Heart Rate, Respiration Rate, SpO<sub>2</sub> | 4 hours |
| Thermoregulation | Temperature | 8 hours |
| Blood Gases | PaO<sub>2</sub>, PaCO<sub>2</sub> | 28 hours |
| Respiration Assistance | IMV, FiO<sub>2</sub> | 12 hours |
| Neurological Observations | GCS, Pupil Size and Reaction | 8 hours |

**Table S4.** Action space discretization grid following [4] and pediatric VIS-based vasopressor intensity [17].

| Discretized Action | IV Fluids Range | Vasopressors Range |
| --- | --- | --- |
| 1 | 0 | 0 |
| 2 | (0.179 – 18.960] | (0.209 – 32.400] |
| 3 | (18.960 – 42.700] | (32.400 – 90.000] |
| 4 | (42.700 – 97.000] | (90.000 – 200.000] |
| 5 | $\geq 97.000$ | $\geq 200.000$ |

## C Extended Results

**Fig S1.**
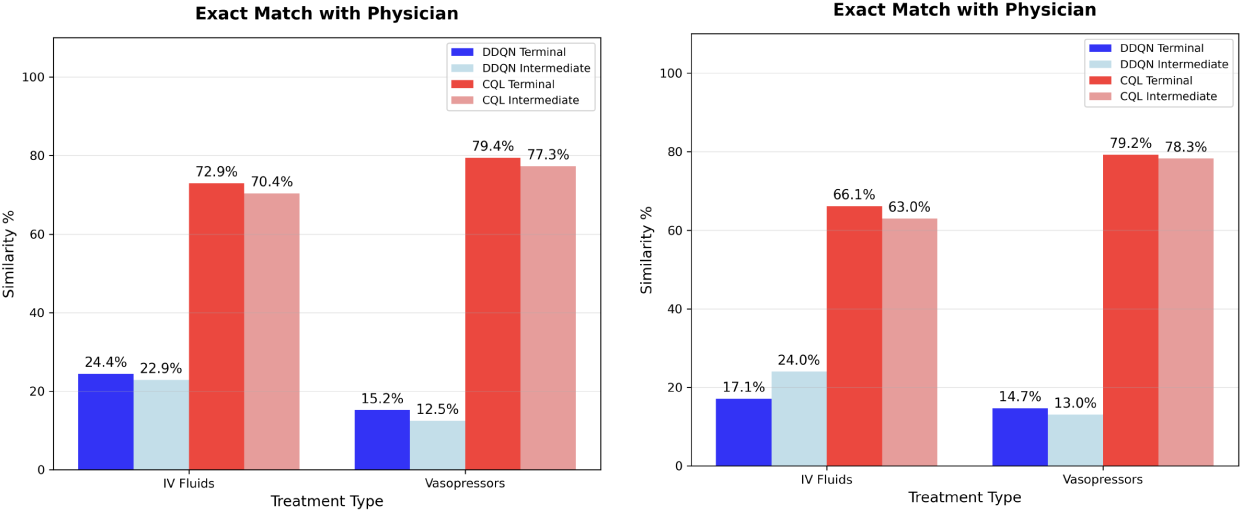
Action similarity at 8-hour and 12-hour temporal resolutions. The relative advantage of CQL over DDQN is preserved.

**Fig S2.**
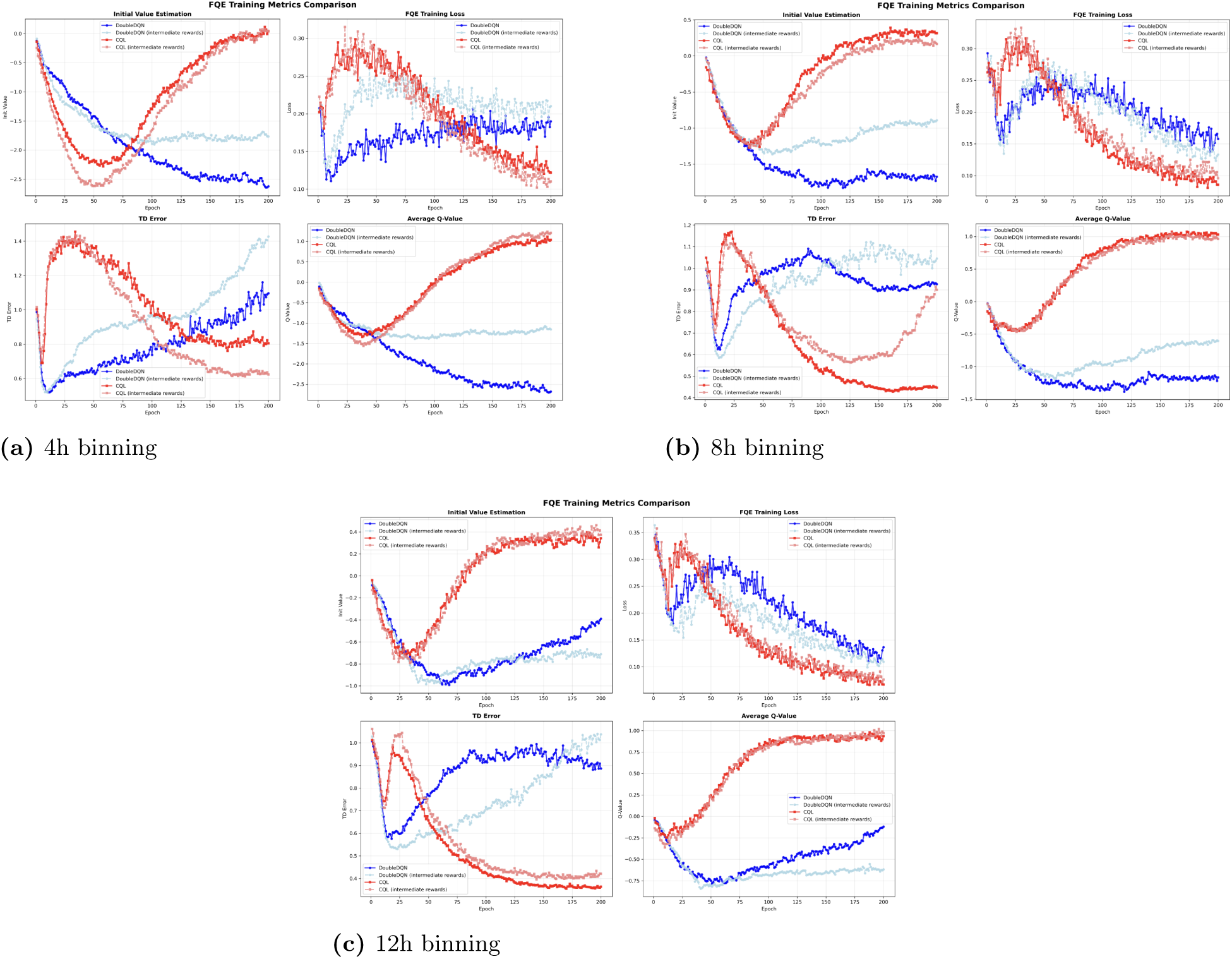
FQE training curves across different temporal resolutions. Two top plots show 4h and 8h side by side; the bottom plot shows 12h binning.

